# The Impact of Agricultural Pesticides on Cancer Incidence in Midwest, United States

**DOI:** 10.1101/2025.06.30.25330578

**Authors:** Benjamin Cox, Brandon M. Varilek, Sushant Mehan, Hossein Moradi Rekabdarkolaee

## Abstract

This analysis examines the relationship between pesticide usage and cancer incidence rates in the Midwest United States using a Bayesian regression model. Pesticide usage data from 1992-2002 and cancer incidence rates from 2017-2022 were analyzed alongside demographic factors such as smoking, binge drinking, and obesity. After selecting the best-fitting distribution, we evaluated the effects of pesticide exposure on cancer rates. The results showed no confident effects from Glyphosate. Atrazine usage results seem to be unreliable, thus no clear conclusion can be made. However, we found Ethalfluralin, 2,4-D, Malathion, and MCPA use was associated with higher overall rates. These findings highlight the need for further research into the potential health risks of pesticide exposure and its role in cancer development.

## 1 Introduction

In the Midwest United States (US), approximately 150 million pounds of agricultural chemicals are used annually (U.S. Geological Survey, 2017). These chemicals include pesticides, herbicides, fungicides, and insecticides, essential chemicals to protect the agricultural industry, one of the largest in the US. Despite the overwhelming use of agricultural chemicals and the mounting evidence of adverse environmental and human health impacts, regulations are lagging behind the extensive use of these chemicals (Ahmad et al., 2024). Although there are examples of harmful chemicals being banned in the US because of the harmful effects on wildlife and humans (e.g., dichloro-diphenyl-trichloroethane [DDT] (Environmental Protection Agency, 2025)). However, there is reluctance to ban other potentially harmful chemicals, many of which still pose significant risk to the health of Americans (U.S. Environmental Protection Agency, 2014; Carozza et al., 2008; Pluth et al., 2019).

Exposure to chemicals used in agriculture pose significant health hazards not only the farmers and agricultural workers in close proximity to the chemicals, but also the general population (Alavanja, 2009; Arcury and Quandt, 1998). The Agricultural Health Study (AHS) has extensively explored the relationship of agricultural chemical exposure farm worker cancer risk, as well as other chronic conditions (e.g., reproductive (Jung et al., 2022), respiratory (Fix et al., 2021; Hoppin et al., 2014), and neurological diseases (Starks et al., 2012; Song et al., 2025)). However, the AHS primarily includes participants from Iowa and North Carolina (Agricultural Health Study, 2025) which limits the usefulness of the results in other agriculturally dense states in the Midwest, such as South Dakota, North Dakota, Nebraska, and Minnesota. Bayesian analyses have not previously. The purpose of this analysis is to use the publicly available datasets and determine the relationship between commonly used agricultural chemicals and cancer incidence rate in Midwest, USA. Bayesian generalized linear model (GLM) is employed for modeling this association.

**Figure 1.**
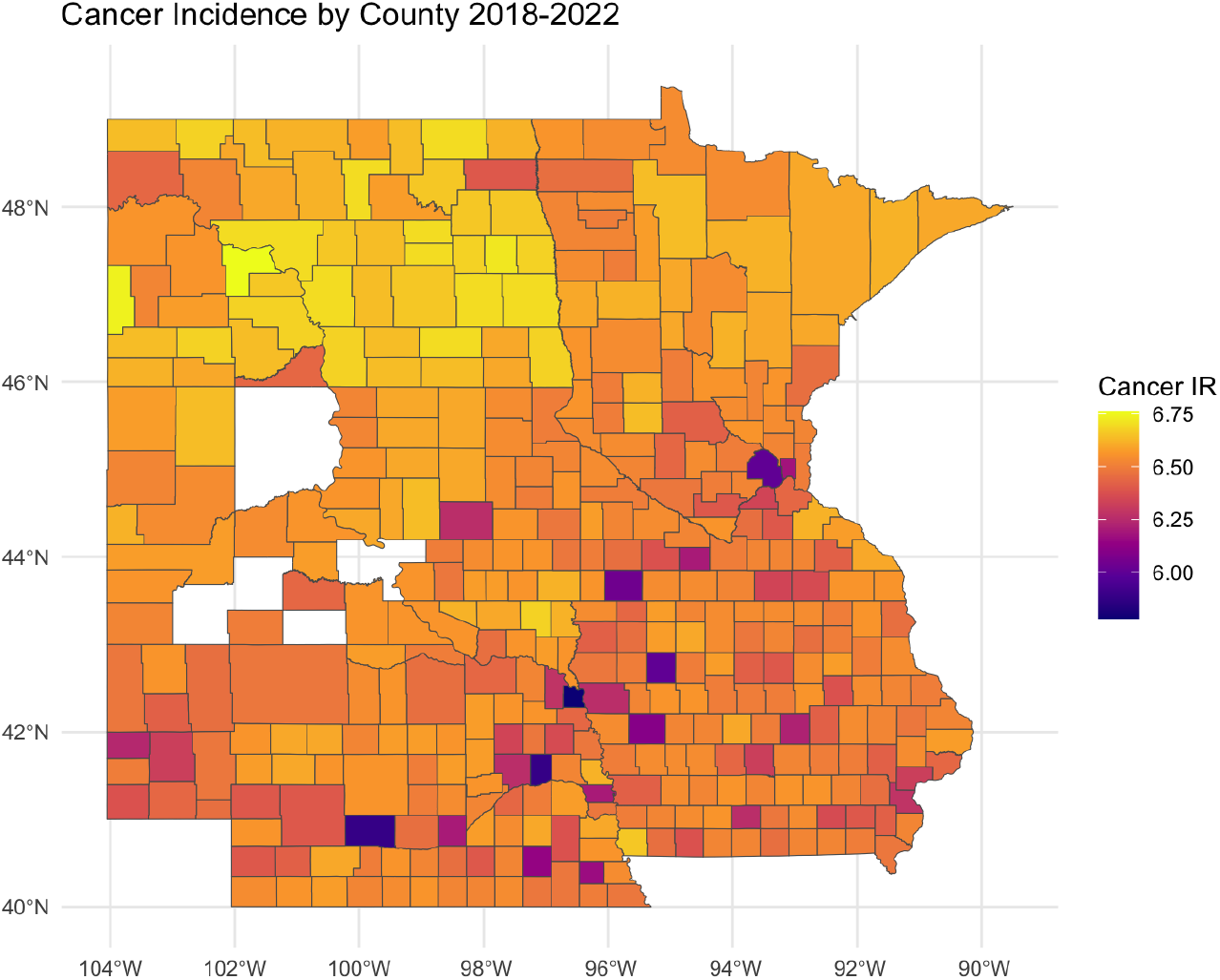
Average incidence rate of cancer by county (2018–2022).

## 2 Data

The data used for analysis is collected various publicly available sources. The pesticide data is provided by the United States Geological Survey (USGS) and was collected as part of the National Water-Quality Assessment project (NAWQA) (U.S. Geological Survey, 2017). The variables obtained from this data for the current study are pesticide compound and low and high estimates of usage. All pesticide usage, measured in kilograms for each compound at the county level and are sorted by Federal Information Processing Standard (FIPS) identification number. This dataset contains almost 400 compounds. However, we only focused on those that are highly used in the Midwest region. These compounds are Glyphosate, Ethalfluralin, Atrazine, 2,4-D, Malathion, MCPA.

The cancer incidence rate along with smoking, drinking, and obesity incidence are obtained from CDC “Local data for better health datasets” (Centers for Disease Control and Prevention, 2018). The dataset provides both the prevalence rate and age-adjusted rates of cancer incidence. This study used the age-adjusted rates since the fluctuation in age could affect the outcome of the models. The demographic data are given as percentages of a total population. Additional demographic variables were obtained from the Agency for Healthcare Research and Quality (AHRQ) datasets for social determinants of health (Agency for Healthcare Research and Quality, 2020).

**Figure 2.**
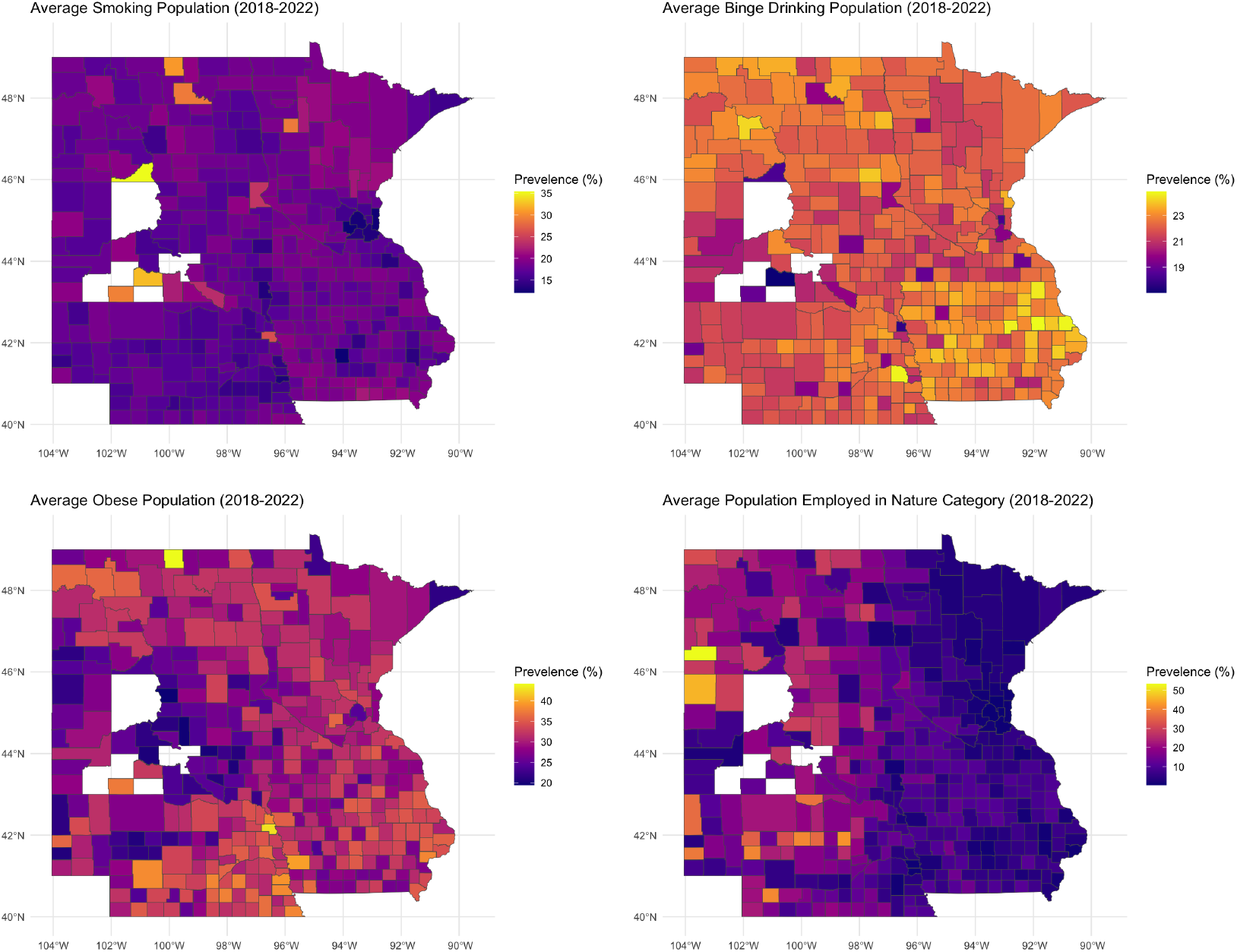
Plots of the smoking rate (top left), binge drinking rate (top right), Obese population (bottom left), and the Nature Workers (bottom right).

**Figure 3.**
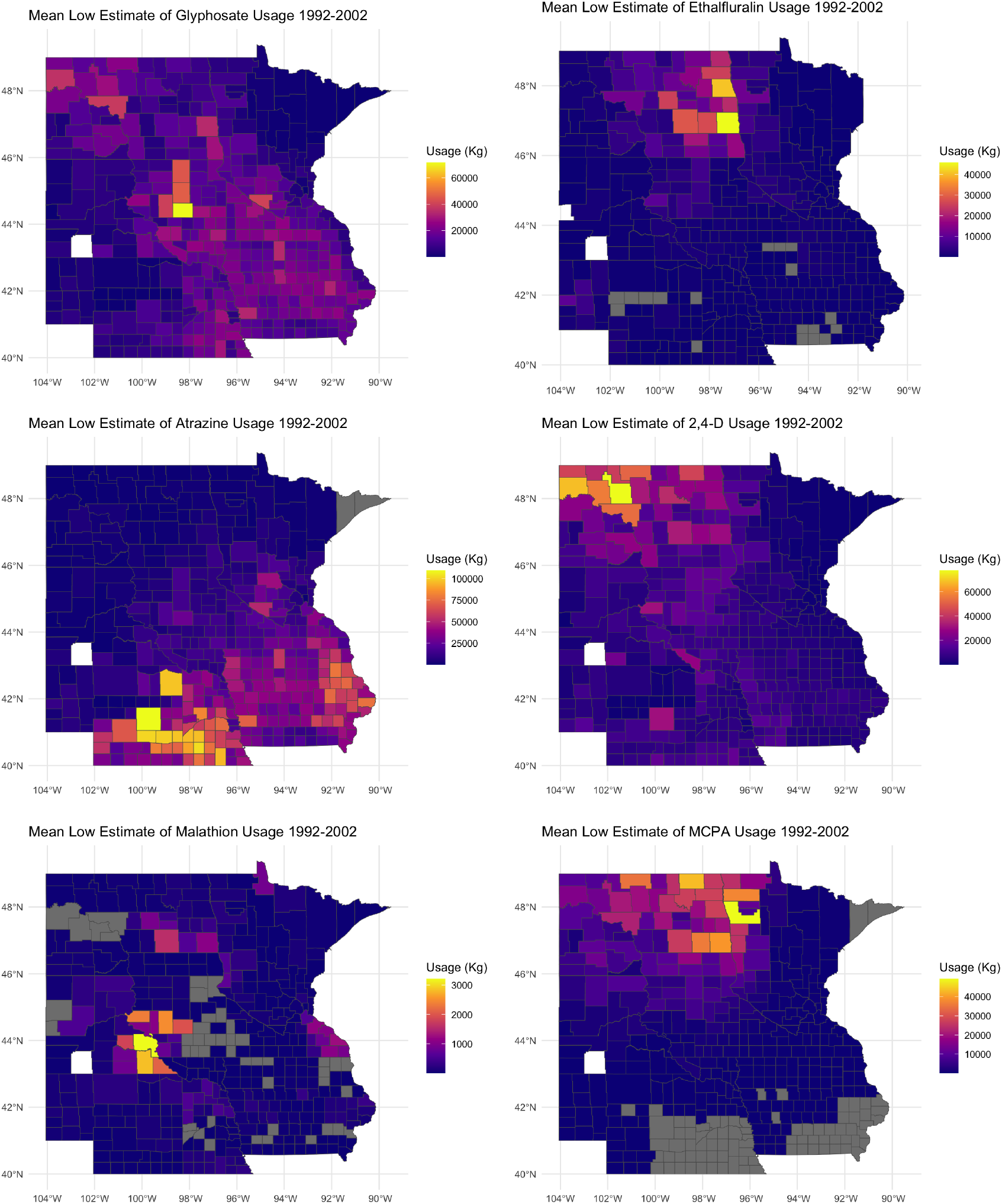
Average glyphosate usage map.

These variables included employed persons working in agriculture, forestry, and mining, Median Household Income, Percent of Uninsured Persons, Total population in poverty, and Total Population. These variables are chosen due to their importance in explaining the cancer incidence and to improve model’s prediction accuracy.

The pesticide usage data was collected from 1992 to 2002. A 10-year county-level average is calculated to identify high- and low-usage regions. The cancer incidence rates were drawn from 2017 to 2022, aggregated as five-year averages. This temporal separation was made to try to account for the known latency period between carcinogen exposure and clinical cancer development, ensuring our analysis captures the likely window for observable population-level effects. Additional demographic variables (2017–2022) were incorporated to control for confounding factors and explain residual variance in incidence rates.

The nature of the data presents a number of challenges. First, the response variable is truncated between 0 and 1 which means standard linear models are not appropriate choice for modeling. Furthermore, there are different sources of uncertainty in the data which needs to be characterize properly. A Bayesian generalized linear model is developed to address these challenges. The model specification is detailed in the next section.

## 3 Methods

To model the effects of multiple predictors on a response variable, a generalized linear model is employed with multiple link. In specific, suppose the response variable **y** = (*y*_1_, …, *y*_*n*_)^*T*^ are observations from an exponential family with mean ***µ*** = (*µ*_1_, …, *µ*_*n*_)^*T*^. Furthermore, suppose that there exists a set of covariates **X** that are related to the mean function through a link function *g*(*·*). This hierarchical model can be written as

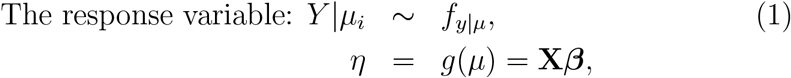

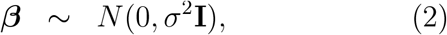

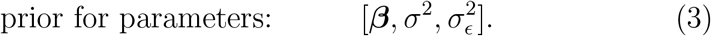

We consider that the parameters are apriori independent, i.e.

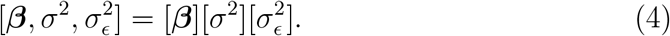

The following distribution functions are used for *f*_*y*|*µ*_(*·*):

1. Normal distribution with random effect for FIPS: *y*_*i*_ *~ N* (*µ*_*i*_, *σ*^2^) with with 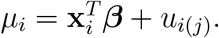.
2. Log-normal distribution with random effect for FIPS: *y*_*i*_ *~ LN* (*µ*_*i*_, *σ*^2^)with 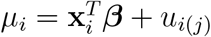
3. Gamma distribution with random effect for FIPS: *y*_*i*_ *~ IG*(*α, γ*_*i*_) with *γ*_*i*_ = *α * µ*_*i*_ where 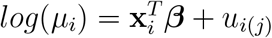.
4. Beta distribution with random effect for FIPS.

The models specifications will be completed by choosing the prior for the parameters. Our general strategy for prior specification is to find proper, yet relatively non-informative, distributions. That is, we seek to specify a prior distribution that spreads its density over a reasonable (practical) range of values while letting the data do most of the informing of the posterior distribution. The coefficients in the mean function is assumed to follow a normal distribution with mean 0 and standard deviation of 10. The random effect for the FIPS, *u*_*i*(*j*)_ is considered to follow a normal distribution with mean 0 and variance 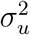 where 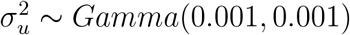.

## 4 Results

All statistical analyses were conducted using R 4.4.1 (R Core Team, 2024). The Bayesian inference was carried out using Markov Chain Monte Carlo (MCMC) techniques to obtain samples from posterior distributions. For model, four separate MCMC chains were run in parallel for 10,000 total iterations. The first 9,000 iterations of each chain were the burn-in iterations and the remaining 1,000 samples used to draw the inference. The `R2JAGS` package (Su and Yajima, 2024) is used to implement normal, log-normal, and Gamma models. The beta-binomial regression models was fitted using the `glmmTMB` package (McGillycuddy et al., 2025). To compare the results of different models, *R*^2^, root mean standard error (RMSE), 10-fold cross validation (CV), and computation time are used. The *R*^2^ and RMSE assess the quality of the fit while the 10-fold CV shows the prediction quality of the model. The user computation time shows the time it takes for a model to be fully executed. The lower values for these criteria shows the better model.

Based on the results in Table 1, the beta-binomial model demonstrates the best overall performance, showing the highest *R*^2^ value along with the lowest RMSE and smallest cross-validation error. While its fast computation time enables more extensive exploratory data analysis, this efficiency may indicate either the model’s relative simplicity or the R package’s optimized implementation. Since it is not common for all theses pesticides to be used simultaneously, the beta-binomial model is used to estimate the effect of pesticide usage individually. In addition to pesticide usage, these models include smoking, binge drinking, obesity prevalence, and nature worker variables.

**Table 1:** Comparison of Bayesian *R*^2^, RMSE, 10-Fold CV, and User Computation Time across different distributions. Models for all distributions run with variables for smoking, drinking, obesity, nature, and all pesticide compounds.

| Dist. | $R^2$ | RMSE [95% CI] | CV | Comp. T. |
| --- | --- | --- | --- | --- |
| Normal | 0.4720 | 0.0043 [0.0021, 0.0079] | 28.36965 | 7.021 |
| LogNormal | 0.5330 | 2.7954 [2.7887, 2.8023] | 354.1332 | 52.736 |
| Gamma | 0.4605 | 2.6649 [2.6586, 2.6713] | 8.4867 | 118.615 |
| Beta-binomial | 0.5590 | 0.00097 [0.0009, 0.0010] | 0.0016 | 1.307 |

Table 2 represents the results of the individual pesticide usage analysis. All these models are based on the beta-binomial regression. This results shows that smoking and binge drinking rates have always positive impact on cancer rates i.e. an increase in smoking and drinking rates leads to an increase in cancer rates. Similar behavior can be seen in nature workers in most cases except for 2,4-D which it seems to have a negative impact. It should be noted that the nature worker is not significant in any of the model in presence of the other variables. The effects of pesticide usage differed across the compounds tested. The effect of Glyphosate use is shown to not to be significant and the effect of Atrazine use results is shown to have negative effect on cancer rates or that Atrazine usage was linked to lower cancer incidence rates. The results shows that Ethalfluralin, 2,4-D, Malathion, and MCPA have a small but positive association with cancer rates. Therefore, suggesting Ethalfluralin, Malathion, MCPA, and 2,4-D usage is associated with higher cancer incidence. The results for other distributions are available in the appendix.

**Table 2:** The *R*^2^, RMSE, regression coefficient estimates and their associated 95% credible intervals in parenthesis for different pesticides.

| Pesticide | Glyphosate | Ethalfuralin | Atrazine | 2,4-D | Malathion | MCPA |
| --- | --- | --- | --- | --- | --- | --- |
| | $R^2 = 0.376$<br>RMSE = 0.001 | $R^2 = 0.464$<br>RMSE = 0.001 | $R^2 = 0.495$<br>RMSE = 0.001 | $R^2 = 0.411$<br>RMSE = 0.001 | $R^2 = 0.439$<br>RMSE = 0.010 | $R^2 = .480$<br>RMSE = 0.001 |
| Variable | **Estimate (95% CI)** |  |  |  |  |  |
| Intercept | -2.85 (-2.93, -2.78) | -2.86 (-2.93, -2.79) | -2.89 (-2.97, -2.82) | -2.85 (-2.92, -2.78) | -2.87 (-2.95, -2.80) | -2.84 (-2.92, -2.76) |
| Smoking | 0.15 (-0.01, 0.31) | 0.17 (0.01, 0.33) | 0.10 (-0.06, 0.26) | 0.16 (0.00, 0.32) | 0.16 (-0.001, 0.32) | 0.17 (-0.001, 0.35) |
| Drinking | 0.83 (0.53, 1.12) | 0.85 (0.56, 1.14) | 0.94 (0.65, 1.24) | 0.83 (0.54, 1.12) | 0.89 (0.58, 1.20) | 0.78 (0.45, 1.11) |
| Obesity | -0.08 (-0.19, 0.03) | -0.09 (-0.19, 0.02) | 0.02 (-0.09, 0.14) | -0.11 (-0.22, -0.01) | -0.07 (-0.18, 0.05) | -0.10 (-0.23, 0.03) |
| Pesticide-Use | 0.00 (-0.03, 0.03) | 0.05 (0.02, 0.07) | -0.04 (-0.05, -0.02) | 0.04 (0.01, 0.07) | 0.04 (0.01, 0.08) | 0.04 (0.02, 0.07) |
| Nat. Workers | 0.04 (-0.03, 0.11) | 0.03 (-0.04, 0.09) | 0.05 (-0.02, 0.11) | 0.00 (-0.08, 0.07) | 0.03 (-0.04, 0.10) | 0.00 (-0.08, 0.08) |

## 5 Discussion

This study investigated the relationship between pesticide usage and cancer incidence rates in the Midwest United States using a beta-binomial regression model to account for demographic, behavioral, and environmental factors. The analysis revealed several important findings that contribute to our understanding of the complex interplay between pesticide exposure and comorbidities on cancer risk.

The model for Ethalfluralin usage shows the strongest indication of an effect on cancer incidence rates with an increase of an estimated 4.9%. The second-strongest association is observed with MCPA usage, which has an average estimated 4.2% increase in cancer incidence rates. However, a key limitation is that MCPA was only used in 307 of 390 counties, so results should be interpreted cautiously. Chemically related to MCPA, 2,4-D exhibits a weaker but still significant association, with an average increase 3.6% in cancer rates. Unlike MCPA, 2,4-D data covers all 390 counties, strengthening the findings. The EPA maintains that 2,4-D poses no carcinogenic risk, but independent studies have limited data linking it to rare cancers like non-hodgekins lymphoma (Burns et al., 2011).While the EPA has not conducted a formal cancer assessment for MCPA (Environmental Protection Agency, n.d.) and regulatory agencies generally deem it non-carcinogenic, recent independent studies challenge this conclusion (von Stackelberg, 2013). Lastly, Malathion shows a slight positive correlation, increasing cancer rates by 4.2% on average, though there was a wide confidence interval for this finding. The EPA highlights ecological risks but asserts no human health concerns (Environmental Protection Agence, 2024). In contrast, the World Health Organization (WHO) classifies malathion as having limited evidence of carcinogenicity in humans and sufficient evidence in animals (World Health Organization, 2016). These contrasting results along with preliminary results from modeling warrant further investigations into cancer risks of exposure to malathion.

Glyphosate did not show a credible interval of association with overall cancer incidence rates. The results mirror results from the EPA which states that no credible evidence exists to support the claim that Glyphosate causes cancer in humans (Environmental Protection Agence, 2025). Alternatively, our results reflected showed that Atrazine exposure was correlated to reduced cancer incidence, despite the known potential effects of Atrazine on human health, such as cancer risk (Wang et al., 2023), endocrine disruption (Wirbisky and Freeman, 2015), and potential birth defects (Krishnapura et al., 2024). Our results may have captured the effects of other predictors and is over confident in its effects. Thus, no strong conclusions can be made on this finding.

Our analysis included other known risk factors that increase cancer risk including smoking (Smolarz et al., 2025; J et al., 2025), alcohol consumption (Jun et al., 2023), and obesity. We found that for every 10% increase in smoking rates with a population, the incidence of cancer increases 1%-1.8% across the models. Interestingly, models including Glyphosate and Atrazine displayed unclear effects of smoking. This result is likely due to issues within modeling, and results should be interpreted with caution. Smoking remains one of the most well-established risk factors for cancer, and its significant impact is clearly reflected in the other models.

For alcohol consumption, the magnitude of the risk increase is amplified. We found a 10% increase of individuals that binge drink within a population was correlated with cancer incidence increasing by 8%-10% across all models. This may be due to the broader impact of alcohol on cancer development across multiple organ systems, whereas smoking is more strongly associated with specific types of cancer (Hydes et al., 2019; Hastings, 2019).

Finally, obesity rates did not show a credible influence on cancer incidence rates in any of the pesticide compound models. We hypothesize that this result may be linked to the reduced life expectancy associated with obesity. Unhealthy lifestyle choices and obesity can shorten life expectancy by years or even decades (Obesity Medicine Association, 2023; Peeters et al., 2003), potentially reducing the likelihood of individuals living to an age where they might be at a higher risk of cancer. However, further research is needed to fully understand the relationship between obesity and cancer outcomes.

## 6 Limitations

A limitation we experienced was absence of additional demographic and environmental variables that could enhance the model’s precision. For example, genetic factors play a significant role in cancer risk, but no publicly available datasets currently provide this information. Similarly, variables such as air pollution, which may contribute to cancer incidence, were not included in our analysis. Incorporating these factors in future research could help explain additional variance and improve the model’s accuracy.

Among the pesticides analyzed in our study, Glyphosate exhibited uncertain effects on cancer incidence rates. Atrazine usage models were overconfident and likely inaccurate. However, we also identified small but statistically significant positive associations between exposure to Ethalfluralin, 2,4-D, Malathion, and MCPA and elevated cancer incidence rates. As previously noted, the EPA has provided limited data on the carcinogenic potential of these pesticides. Our findings highlight the need for further research and stricter regulatory standards for pesticide use in the United States.

However, one limitation of our analysis is the lack of detailed data on specific cancer site incidence rates at the county level. This data is not publicly available due to data suppression for smaller populations and anonymity concerns. As discussed, many of the compounds selected have limited research suggesting they may be linked to specific types of cancer. By using general cancer incidence rates, we may miss the potential carcinogenic effects of these compounds on particular cancer sites. Future research could focus on analyzing site-specific cancer rates to better understand the relationship between pesticide exposure and specific types of cancer.

A critical assumption in our study is the latency period between pesticide exposure and the development of cancer. While we selected pesticide usage data from 1992–2002 and cancer incidence data from 2017–2022 to account for a 10-to 20-year latency period, this assumption may not hold for all pesticides or cancer types. Pesticides have been used since the early 1900s, and exposure outside the selected time frame could also influence cancer rates. Future research could explore the effects of pesticide usage over longer time periods and refine our understanding of latency periods.

Finally, as our study is confined only to the Midwest region, including South Dakota, North Dakota, Nebraska, Minnesota, and Iowa. While this region was chosen for its consistent agricultural practices, demographics, and health outcomes, it limits the generalization of our findings. After excluding certain counties, our sample size was reduced to 390, which is relatively small. A larger sample size, or data at a more granular level (e.g., sub-county or zip code), could improve the model’s predictive power and provide more nuanced insights.

## 7 Conclusion

In conclusion, the results consistently demonstrated strong positive associations between smoking, alcohol consumption, and their relationship to cancer incidence rates. We aimed to provided valuable insights on the relationship between pesticide usage and cancer incidence rates in the Midwest. These findings highlight the importance of addressing known risk factors, such as smoking and binge drinking, while also calling for further investigation into the health effects of specific pesticides. By addressing the limitations of this research and expanding its scope, future studies can build on these findings to inform public health policies and protect vulnerable rural populations from the potential risks of pesticide exposure.

## Data Availability

All the websites for data are available in the manuscript along with the date that it was accessed.

## 8 Appendix: The results for other models

### Normal Distribution Results

**Table 3:** Normal distribution estimates and 95% confidence intervals using Glyphosate usage, *R*^2^ = 0.4669, RMSE = 0.0032.

| Variable | Estimate | 95% CI Lower | 95% CI Upper |
| --- | --- | --- | --- |
| Intercept | 0.055 | 0.025 | 0.085 |
| Smoking | 0.005 | -0.041 | 0.051 |
| Binge Drinking | 0.047 | -0.066 | 0.160 |
| Obesity Prevalence | -0.004 | -0.037 | 0.030 |
| Nature Workers | 0.003 | -0.013 | 0.019 |
| Pesticide Usage | -0.0005 | -0.011 | 0.010 |

**Table 4:** Normal distribution estimates and 95% confidence intervals using Ethalfluralin usage, *R*^2^ = 0.4683, RMSE = 0.0031.

| Variable | Estimate | 95% CI Lower | 95% CI Upper |
| --- | --- | --- | --- |
| Intercept | 0.057 | 0.032 | 0.082 |
| Smoking | 0.005 | -0.038 | 0.047 |
| Binge Drinking | 0.038 | -0.056 | 0.132 |
| Obesity Prevalence | -0.006 | -0.038 | 0.027 |
| Nature Workers | 0.002 | -0.014 | 0.018 |
| Pesticide Usage | 0.003 | -0.009 | 0.015 |

**Table 5:** Normal distribution estimates and 95% confidence intervals using Atrazine usage, *R*^2^ = 0.4628, RMSE = 0.0032.

| Variable | Estimate | 95% CI Lower | 95% CI Upper |
| --- | --- | --- | --- |
| Intercept | 0.063 | 0.032 | 0.095 |
| Smoking | -0.001 | -0.049 | 0.046 |
| Binge Drinking | 0.016 | -0.102 | 0.134 |
| Obesity Prevalence | -0.006 | -0.042 | 0.031 |
| Nature Workers | 0.002 | -0.014 | 0.018 |
| Pesticide Usage | -0.001 | -0.008 | 0.006 |

**Table 6:** Normal distribution estimates and 95% confidence intervals using 2,4-D usage, *R*^2^ = 0.4656, RMSE = 0.0033.

| Variable | Estimate | 95% CI Lower | 95% CI Upper |
| --- | --- | --- | --- |
| Intercept | 0.055 | 0.022 | 0.088 |
| Smoking | 0.006 | -0.041 | 0.053 |
| Binge Drinking | 0.045 | -0.075 | 0.165 |
| Obesity Prevalence | -0.005 | -0.041 | 0.031 |
| Nature Workers | 0.002 | -0.015 | 0.019 |
| Pesticide Usage | 0.002 | -0.009 | 0.013 |

**Table 7:**
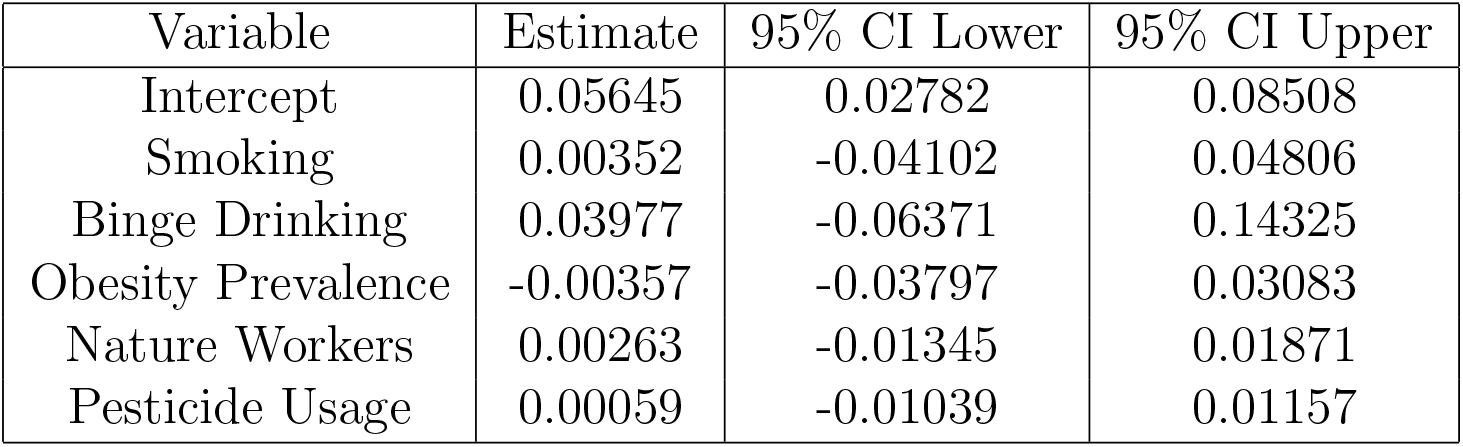
Normal distribution estimates and 95% confidence intervals using Malathion usage, *R*^2^ = 0.4634, RMSE = 0.0032.

**Table 8:** Normal distribution estimates and 95% confidence intervals using MCPA usage, *R*^2^ = 0.4664, RMSE = 0.0032.

| Variable | Estimate | 95% CI Lower | 95% CI Upper |
| --- | --- | --- | --- |
| Intercept | 0.063 | 0.031 | 0.095 |
| Smoking | 0.001 | -0.043 | 0.045 |
| Binge Drinking | 0.018 | -0.098 | 0.135 |
| Obesity Prevalence | -0.008 | -0.041 | 0.025 |
| Nature Workers | 0.001 | -0.015 | 0.017 |
| Pesticide Usage | 0.002 | -0.006 | 0.011 |

### Log-Normal Distribution Results

**Table 9:** Log-Normal distribution estimates and 95% confidence intervals using Glyphosate usage, *R*^2^ = 0.5330, RMSE = 2.7965.

| Variable | Estimate | 95% CI Lower | 95% CI Upper |
| --- | --- | --- | --- |
| Intercept | -2.845 | -2.878 | -2.812 |
| Smoking | 0.066 | 0.014 | 0.119 |
| Binge Drinking | 0.555 | 0.435 | 0.676 |
| Obesity Prevalence | -0.081 | -0.121 | -0.041 |
| Nature Workers | 0.036 | 0.017 | 0.055 |
| Pesticide Usage | -0.004 | -0.016 | 0.008 |

**Table 10:** Log-Normal distribution estimates and 95% confidence intervals using Ethalfluralin usage, *R*^2^ = 0.5330, RMSE = 2.7965.

| Variable | Estimate | 95% CI Lower | 95% CI Upper |
| --- | --- | --- | --- |
| Intercept | -2.840 | -2.873 | -2.806 |
| Smoking | 0.072 | 0.019 | 0.124 |
| Binge Drinking | 0.544 | 0.424 | 0.665 |
| Obesity Prevalence | -0.100 | -0.138 | -0.062 |
| Nature Workers | 0.027 | 0.008 | 0.046 |
| Pesticide Usage | 0.045 | 0.031 | 0.058 |

**Table 11:** Log-Normal distribution estimates and 95% confidence intervals using Atrazine usage, *R*^2^ = 0.5338, RMSE = 2.7965.

| Variable | Estimate | 95% CI Lower | 95% CI Upper |
| --- | --- | --- | --- |
| Intercept | -2.864 | -2.893 | -2.835 |
| Smoking | 0.031 | -0.022 | 0.083 |
| Binge Drinking | 0.619 | 0.511 | 0.727 |
| Obesity Prevalence | -0.025 | -0.067 | 0.016 |
| Nature Workers | 0.033 | 0.014 | 0.052 |
| Pesticide Usage | -0.026 | -0.034 | -0.018 |

**Table 12:** Log-Normal distribution estimates and 95% confidence intervals using 2,4-D usage, *R*^2^ = 0.5327, RMSE = 2.7965.

| Variable | Estimate | 95% CI Lower | 95% CI Upper |
| --- | --- | --- | --- |
| Intercept | -2.826 | -2.861 | -2.791 |
| Smoking | 0.057 | 0.004 | 0.111 |
| Binge Drinking | 0.496 | 0.366 | 0.625 |
| Obesity Prevalence | -0.103 | -0.141 | -0.066 |
| Nature Workers | 0.022 | 0.002 | 0.042 |
| Pesticide Usage | 0.029 | 0.016 | 0.042 |

**Table 13:** Log-Normal distribution estimates and 95% confidence intervals using Malathion usage, *R*^2^ = 0.5329, RMSE = 2.7965.

| Variable | Estimate | 95% CI Lower | 95% CI Upper |
| --- | --- | --- | --- |
| Intercept | -2.860 | -2.897 | -2.823 |
| Smoking | 0.078 | 0.025 | 0.131 |
| Binge Drinking | 0.602 | 0.467 | 0.736 |
| Obesity Prevalence | -0.076 | -0.115 | -0.037 |
| Nature Workers | 0.036 | 0.017 | 0.055 |
| Pesticide Usage | 0.009 | -0.004 | 0.022 |

**Table 14:** Log-Normal distribution estimates and 95% confidence intervals using MCPA usage, *R*^2^ = 0.5325, RMSE = 2.7965.

| Variable | Estimate | 95% CI Lower | 95% CI Upper |
| --- | --- | --- | --- |
| Intercept | -2.841 | -2.875 | -2.807 |
| Smoking | 0.069 | 0.018 | 0.120 |
| Binge Drinking | 0.550 | 0.425 | 0.676 |
| Obesity Prevalence | -0.099 | -0.137 | -0.060 |
| Nature Workers | 0.023 | 0.004 | 0.042 |
| Pesticide Usage | 0.034 | 0.023 | 0.044 |

### Gamma Distribution Results

**Table 15:** Inverse-Gamma distribution estimates and 95% confidence intervals using Glyphosate usage, *R*^2^ = 0.4622, RMSE = 2.6661.

| Variable | Estimate | 95% CI Lower | 95% CI Upper |
| --- | --- | --- | --- |
| Intercept | 2.854 | 2.819 | 2.888 |
| Smoking | -0.072 | -0.123 | -0.022 |
| Binge Drinking | -0.586 | -0.708 | -0.464 |
| Obesity Prevalence | 0.078 | 0.040 | 0.116 |
| Nature Workers | -0.035 | -0.054 | -0.017 |
| Pesticide Usage | 0.005 | -0.007 | 0.016 |

**Table 16:** Inverse-Gamma distribution estimates and 95% confidence intervals using Ethalfluralin usage, *R*^2^ = 0.4643, RMSE = 2.6661.

| Variable | Estimate | 95% CI Lower | 95% CI Upper |
| --- | --- | --- | --- |
| Intercept | 2.852 | 2.820 | 2.884 |
| Smoking | -0.079 | -0.128 | -0.029 |
| Binge Drinking | -0.584 | -0.703 | -0.465 |
| Obesity Prevalence | 0.094 | 0.058 | 0.130 |
| Nature Workers | -0.028 | -0.046 | -0.010 |
| Pesticide Usage | -0.044 | -0.057 | -0.032 |

**Table 17:** Inverse-Gamma distribution estimates and 95% confidence intervals using Atrazine usage, *R*^2^ = 0.4633, RMSE = 2.6661.

| Variable | Estimate | 95% CI Lower | 95% CI Upper |
| --- | --- | --- | --- |
| Intercept | 2.862 | 2.829 | 2.896 |
| Smoking | -0.030 | -0.080 | 0.020 |
| Binge Drinking | -0.614 | -0.729 | -0.499 |
| Obesity Prevalence | 0.027 | -0.012 | 0.066 |
| Nature Workers | -0.032 | -0.050 | -0.015 |
| Pesticide Usage | 0.026 | 0.018 | 0.034 |

**Table 18:** Inverse-Gamma distribution estimates and 95% confidence intervals using 2,4-D usage, *R*^2^ = 0.4641, RMSE = 2.6661.

| Variable | Estimate | 95% CI Lower | 95% CI Upper |
| --- | --- | --- | --- |
| Intercept | 2.849 | 2.816 | 2.881 |
| Smoking | -0.074 | -0.124 | -0.024 |
| Binge Drinking | -0.581 | -0.701 | -0.461 |
| Obesity Prevalence | 0.101 | 0.065 | 0.137 |
| Nature Workers | -0.022 | -0.040 | -0.003 |
| Pesticide Usage | -0.028 | -0.040 | -0.016 |

**Table 19:** Inverse-Gamma distribution estimates and 95% confidence intervals using Malathion usage, *R*^2^ = 0.4627, RMSE = 2.6661.

| Variable | Estimate | 95% CI Lower | 95% CI Upper |
| --- | --- | --- | --- |
| Intercept | 2.856 | 2.820 | 2.892 |
| Smoking | -0.072 | -0.124 | -0.020 |
| Binge Drinking | -0.599 | -0.731 | -0.467 |
| Obesity Prevalence | 0.082 | 0.044 | 0.121 |
| Nature Workers | -0.034 | -0.052 | -0.016 |
| Pesticide Usage | -0.009 | -0.021 | 0.003 |

**Table 20:** Inverse-Gamma distribution estimates and 95% confidence intervals using MCPA usage, *R*^2^ = 0.4646, RMSE = 2.6661.

| Variable | Estimate | 95% CI Lower | 95% CI Upper |
| --- | --- | --- | --- |
| Intercept | 2.840 | 2.806 | 2.873 |
| Smoking | -0.062 | -0.110 | -0.014 |
| Binge Drinking | -0.547 | -0.670 | -0.425 |
| Obesity Prevalence | 0.095 | 0.058 | 0.132 |
| Nature Workers | -0.023 | -0.041 | -0.005 |
| Pesticide Usage | -0.034 | -0.044 | -0.024 |

## References

Agency for Healthcare Research and Quality. Social determinants of health database (beta version), 2020. URL https://www.ahrq.gov/data/index.html. Accessed: 2025-02-27.

Agricultural Health Study. About the study, 2025. URL https://aghealth.nih.gov/about/. Accessed: 2025-06-18.

Md Faruque Ahmad, Fakhruddin Ali Ahmad, Abdulrahman A. Alsayegh, Md. Zeyaullah, Abdullah M. AlShahrani, Khursheed Muzammil, Abdullah Ali Saati, Shadma Wahab, Ehab Y. Elbendary, Nahla Kambal, Mohamed H. Abdelrahman, and Sohail Hussain. Pesticides impacts on human health and the environment with their mechanisms of action and possible countermeasures. Heliyon, 10(7):e29128, 2024. ISSN 2405-8440. doi: 10.1016/j.heliyon.2024.e29128. URL https://www.sciencedirect.com/science/article/pii/S2405844024051594.

MCR Alavanja. Introduction: Pesticides use and exposure, extensive world-wide. Reviews on Environmental Health, 2009. doi: 10.1515/REVEH.2009.24.4.303.

TA Arcury and S.A. Quandt. Chronic agricultural chemical exposure among migrant and seasonal farmworkers. Society Natural Resources, 1998. doi: 10.1080/08941929809381121.

Carol Burns, Kenneth Bodner, Gerard Swaen, James Collins, Kathy Beard, and Marcia Lee. Cancer incidence of 2, 4-d production workers. International Journal of Environmental Research and Public Health, 8(9):3579–3590, 2011.

Susan E. Carozza, Bo Li, Kai Elgethun, and Ryan Whitworth. Risk of childhood cancers associated with residence in agriculturally intense areas in the united states. Environmental Health Perspectives, 2008. doi: 10.1289/ehp.9967.

Centers for Disease Control and Prevention. Places: Local data for better health, county data 2019–2024 release, 2018. URL https://www.cdc.gov/places. Accessed: 2025-02-27.

Environmental Protection Agence. Epa releases proposed protections for pesticide malathion, 2024. URL https://www.epa.gov/pesticides/epa-releases-proposed-protections-pesticide-malathion. Accessed: 2025-06-23.

Environmental Protection Agence. Glyphosate, 2025. URL https://www.epa.gov/ingredients-used-pesticide-products/glyphosate. Accessed: 2025-06-18.

Environmental Protection Agency. Ddt - a brief history and status, 2025. URL https://www.epa.gov/ingredients-used-pesticide-products/ddt-brief-history-and-status.

Environmental Protection Agency. 2-methyl-4-chlorophenoxyacetic acid (mcpa), n.d. URL https://iris.epa.gov/ChemicalLanding/&substance_nmbr=66. Accessed: 2025-06-18.

Jonathan Fix, Isabella Annesi-Maesano, Isabelle Baldi, Mathilde Boulanger, Soo Cheng, Sandra Cortes, Jean-Charles Dalphin, Mohamed Aqiel Dalvie, Bruno Degano, Jeroen Douwes, Wijnand Eduard, Grethe Elholm, Catterina Ferreccio, Anne-Helen Harding, Mohamed Jeebhay, Kevin M Kelly, Hans Kromhout, Ewan MacFarlane, Cara Nichole Maesano, Diane Catherine Mitchell, Hussein Mwanga, Saloshni Naidoo, Beyene Negatu, Dorothy Ngajilo, Karl-Christian Nordby, Christine G Parks, Marc B Schenker, Aesun Shin, Torben Sigsgaard, Malcolm Sim, Thibaud Soumagne, Peter Thorne, Keun-Young Yoo, and Jane A Hoppin and. Gender differences in respiratory health outcomes among farming cohorts around the globe: findings from the agricoh consortium. Journal of Agromedicine, 26(2): 97–108, 2021. doi: 10.1080/1059924X.2020.1713274.

C Hastings. When it comes to cancer, how does alcohol compare to smoking?, 2019. URL https://www.icr.ac.uk/research-and-discoveries/cancer-blogs/detail/science-talk/when-it-comes-to-cancer-how-does-alcohol-compare-to-smoking. Accessed: 2025-06-24.

Jane A Hoppin, David M Umbach, Stuart Long, Jessica L Rinsky, Paul K Henneberger, Paivi M Salo, Darryl C Zeldin, Stephanie J London, Michael C R Alavanja, Aaron Blair, Laura E Beane Freeman, and Dale P Sandler. Respiratory disease in united states farmers. Occupational and Environmental Medicine, 71(7):484–491, 2014. doi: 10.1136/oemed-2013-101983.

Theresa J. Hydes, Robyn Burton, Hazel Inskip, Mark A. Bellis, and Nick Sheron. A comparison of gender-linked population cancer risks between alcohol and tobacco: how many cigarettes are there in a bottle of wine? BMC Public Health, 19, 2019. doi: 10.1186/s12889-019-6576-9.

Hovanec J, Kendzia B, Olsson A, Schüz J, Kromhout H, Vermeulen R, Peters S, Gustavsson P, Migliore E, Radoi L, Barul C, Consonni D, Caporaso NE, Landi MT, Field JK, Karrasch S, Wichmann HE, Siemiatycki J, Parent ME, Richiardi L, Simonato L, Jöckel KH, Ahrens W, Pohlabeln H, Fernández-Tardón G, Zaridze D, McLaughlin JR, Demers PA, Światkowska B, Lissowska J, Pándics T, Fabianova E, Mates D, Schejbalova M, Foretova L, Janout V, Boffetta P, Forastiere F, Straif K, Brüning T, and Behrens T. Socioeconomic status, smoking, and lung cancer: Mediation and bias analysis in the synergy study. Epidemiology, 36:245–252, 2025. doi: 10.1097/EDE.0000000000001807.

S. Jun, H. Park, U. J. Kim, E. J. Choi, H. A. Lee, B. Park, S. Y. Lee, S. H. Jee, and H. Park. Cancer risk based on alcohol consumption levels: a comprehensive systematic review and meta-analysis. Epidemiology and Health, 45, 2023. doi: 10.4178/epih.e2023092.

Audrey Y Jung, Thomas U Ahearn, Sabine Behrens, Pooja Middha, Manjeet K Bolla, Qin Wang, Volker Arndt, Kristan J Aronson, Annelie Augustinsson, Laura E Beane Freeman, Heiko Becher, Hermann Brenner, Federico Canzian, Lisa A Carey, CTS Consortium, Kamila Czene, A Heather Eliassen, Mikael Eriksson, D Gareth Evans, Jonine D Figueroa, Lin Fritschi, Marike Gabrielson, Graham G Giles, Pascal Guénel, Andreas Hadjisavvas, Christopher A Haiman, Niclas Håkansson, Per Hall, Ute Hamann, Reiner Hoppe, John L Hopper, Anthony Howell, David J Hunter, Anika Hüsing, Rudolf Kaaks, Veli-Matti Kosma, Stella Koutros, Peter Kraft, James V Lacey, Loic Le Marchand, Jolanta Lissowska, Maria A Loizidou, Arto Mannermaa, Tabea Maurer, Rachel A Murphy, Andrew F Olshan, Håkan Olsson, Alpa V Patel, Charles M Perou, Gad Rennert, Rana Shibli, Xiao-Ou Shu, Melissa C Southey, Jennifer Stone, Rulla M Tamimi, Lauren R Teras, Melissa A Troester, Thérèse Truong, Celine M Vachon, Sophia S Wang, Alicja Wolk, Anna H Wu, Xiaohong R Yang, Wei Zheng, Alison M Dunning, Paul D P Pharoah, Douglas F Easton, Roger L Milne, Nilanjan Chatterjee, Marjanka K Schmidt, Montserrat García-Closas, and Jenny Chang-Claude. Distinct reproductive risk profiles for intrinsic-like breast cancer subtypes: Pooled analysis of population-based studies. Journal of the National Cancer Institute, 114(12):1706–1719, 06 2022. doi: 10.1093/jnci/djac117.

Sunaya R. Krishnapura, Elizabeth McNeer, William D. Dupont, and Stephen W. Patrick. County-level atrazine use and gastroschisis. JAMA Network Open, 7(5):e2410056–e2410056, 05 2024. doi: 10.1001/jamanetworkopen.2024.10056.

Maeve McGillycuddy, Gordana Popovic, Benjamin M Bolker, and David I Warton. Parsimoniously fitting large multivariate random effects in glmmtmb. Journal of Statistical Software, 112:1–19, 2025.

Obesity Medicine Association. Obesity and life expectancy trends in the u.s., 2023. URL https://obesitymedicine.org/blog/obesity-and-life-expectancy-trends-in-the-u-s/. Accessed: 2025-06-24.

Anna Peeters, Jan J Barendregt, Frans Willekens, Johan P Mackenbach, Abdullah Al Mamun, Luc Bonneux, and NEDCOM, the Netherlands Epidemiology and Demography Compression of Morbidity Research Group. Obesity in adulthood and its consequences for life expectancy: A life-table analysis. Annals of Internal Medicine, 138(1):24–32, 2003. doi: 10.7326/0003-4819-138-1-200301070-00008.

Thaís Bremm Pluth, Lucas Adalberto Geraldi Zanini, and Iara Denise Endruweit Battisti. Pesticide exposure and cancer: An integrative literature review. Saúde Em Debate, 2019. doi: 10.1590/0103-1104201912220.

R Core Team. R: A Language and Environment for Statistical Computing. R Foundation for Statistical Computing, Vienna, Austria, 2024. URL https://www.R-project.org/.

Beata Smolarz, Honorata Lukasiewicz, Dariusz Samulak, Ewa Piekarska, Rados-law Kolaciński, and Hanna Romanowicz. Lung cancer—epidemiology, pathogenesis, treatment and molecular aspect (review of literature). International Journal of Molecular Sciences, 26(5), 2025. doi: 10.3390/ijms26052049.

Shengfang Song, Zhehui Luo, Brenda L. Plassman, Xuemei Huang, Yaqun Yuan, Srishti Shrestha, Christine G. Parks, Jonathan N. Hofmann, Laura E. Beane Freeman, Dale P. Sandler, and Honglei Chen. Self-reported motor and nonmotor symptoms, prodromal parkinson’s disease probability, and incident parkinson’s disease in us farmers. Movement Disorders, 40(5):855–868, 2025. doi: 10.1002/mds.30149.

Sarah E. Starks, Fred Gerr, Freya Kamel, Charles F. Lynch, Michael P. Jones, Michael C. Alavanja, Dale P. Sandler, and Jane A. Hoppin. Neurobehavioral function and organophosphate insecticide use among pesticide applicators in the agricultural health study. Neurotoxicology and Teratology, 34 (1):168–176, 2012. doi: 10.1016/j.ntt.2011.08.014.

Yu-Sung Su and Masanao Yajima. R2jags: Using R to Run ‘JAGS’, 2024. URL https://CRAN.R-project.org/package=R2jags. R package version 0.8-9.

U.S. Environmental Protection Agency. DDT - a brief history and status, 2014. URLhttps://www.epa.gov/ingredients-used-pesticide-products/ddt-brief-history-and-status.

U.S. Geological Survey. Pesticides prevalent in Midwestern streams, 2017. URL https://www.usgs.gov/news/national-news-release/pesticides-prevalent-midwestern. Accessed: 2025-03-01.

Katherine von Stackelberg. A systematic review of carcinogenic outcomes and potential mechanisms from exposure to 2, 4-d and mcpa in the environment. Journal of toxicology, 2013(1):371610, 2013.

Mengqi Wang, Junyu Chen, Shuhua Zhao, Jingying Zheng, Kang He, Wei Liu, Weixin Zhao, Jingze Li, Kai Wang, Yuru Wang, Jian Liu, and Lijing Zhao. Atrazine promotes breast cancer development by suppressing immune function and upregulating mmp expression. Ecotoxicology and Environmental Safety, 253:114691, 2023. ISSN 0147-6513. doi: 10.1016/j.ecoenv.2023.114691. URL https://www.sciencedirect.com/science/article/pii/S0147651323001951.

Sara E. Wirbisky and Jennifer L. Freeman. Atrazine exposure and reproductive dysfunction through the hypothalamus-pituitary-gonadal (hpg) axis. Toxics, 3(4):414–450, 2015. doi: 10.3390/toxics3040414. URL https://www.mdpi.com/2305-6304/3/4/414.

World Health Organization. Use of malathion for vector control. report of a who meeting, geneva, 16–17 may 2016, 2016. URL https://www.who.int/publications/i/item/9789241510578. Accessed: 2025-06-19.

